# Analytical approaches for medication reconciliation-related topics: a scoping review

**DOI:** 10.1101/2025.03.16.25324060

**Authors:** Xinyu Yao, Amogh Ananda Rao, Rema Padman

## Abstract

**Objective:** This scoping review examines literature related to analytical methods for medication reconciliation in the digital era, particularly using artificial intelligence and operations research approaches, and analyzes their effectiveness in reducing medication errors and improving the accuracy of medication lists during care transitions.

**Materials and Methods:** Following PRISMA-ScR guidelines, we performed a comprehensive literature search in PubMed, Web of Science, ACM, INFORMS, IEEE, and CINAHL databases for English-language studies until December 2023 that explored artificial intelligence, machine learning, and operations research methods for medication reconciliation.

**Results:** We identified 64 unique studies that are closely related to our research topic, with 53% published since 2020 and 27% U.S.-based. Only 8% directly addressed the complete medication reconciliation process; the remainder focused on related areas, including adverse drug event detection/prediction and medication error detection. Merely 7 studies used decision-theoretic operations research methods, while most used machine learning models and only 5 studies used a combination of artificial intelligence and operations research methods for general medication reconciliation purposes.

**Conclusions:** The reviewed literature provides growing evidence of research on adverse event detection for a single drug type but limited work on investigating the holistic incomplete/inaccurate list of prescribed medications for a patient. We also found that most of the literature focused on single methodologies for medication reconciliation. Future studies need to explore how to leverage predictive, prescriptive, and generative analytics, combining both artificial intelligence, including machine learning and generative AI, and operations research approaches to improve medication reconciliation for care transition safety with medication management.

## Introduction

In the increasingly fast-paced and complex healthcare delivery systems today, medication errors are a persistent challenge, leading to serious adverse consequences for the patient, including adverse drug events (ADE), increased hospitalizations, and higher healthcare costs. [1] Improper medication-related injuries harm more than 1.3 million people every year in the US alone. [2] This issue is particularly concerning for patients with polypharmacy, defined as the use of five or more medications, which is associated with an elevated risk of negative care outcomes. [3, 4] A harmful cycle known as prescribing cascades can occur when a patient’s side effects from their existing medications are misdiagnosed as a new medical condition, leading to the addition of another, potentially avoidable, medication. [5] Along with prescribing cascades, poor adherence to medication and transitions between home and healthcare facilities further increase the risk of medication errors and adverse events. [6] Therefore, effective medication error management is crucial for ensuring patient safety and improving overall health outcomes. [7–9]

Among medication management strategies, the Joint Commission has identified medication reconciliation as a key patient safety practice to reduce medication errors, a critical challenge in patient management during care transitions. [10] Medication reconciliation is a critical step that is designed to create and verify an accurate and complete patient medication list at every transition point in the care pathway. [10–12] Formally, medication reconciliation consists of 1) developing a list of current medications based on admission, discharge and transfer (ADT) orders across the patient’s previous care encounters and hospitalizations; 2) developing a list of medications to be prescribed based on the new diagnosis; 3) comparing the medications on the two lists; 4) making clinical decisions for medication adjustment based on the comparison; and 5) communicating the new list to appropriate caregivers and to the patient. [10, 13] This process aims to resolve both clerical errors, such as omissions or duplications on the list, and clinical issues, such as under/over-treatment or prescribing combinations of drugs with known interactions. By ensuring the patient’s existing and newly adjusted medications are compatible and can effectively address all medical conditions, medication reconciliation reduces the risk of ADEs and under/over-treatments. Thus, it plays a crucial role in enhancing patient safety, minimizing potential medication errors and ADEs throughout their healthcare journey, and improving care quality and health outcomes. [14, 15]

Despite ongoing efforts to implement medication reconciliation, challenges persist in realizing its full effectiveness in practice. [16] Errors and omissions that occur during the reconciliation process can even propagate through subsequent stages, including prescription, dispensing, administration, and monitoring. [17] Traditional medication reconciliation requires the manual collection and review of medication lists from prescribers, patients, and data systems, making it time-consuming. [18] Efforts to improve this process have primarily focused on enhancing workflow tasks and integrating IT solutions to ensure accurate documentation and smooth transitions between care settings. [4, 19] However, these process-oriented tasks face continuing challenges with low consistency and standardization of application, fallible memories of patients, incomplete or inaccurate medical histories, poor collaboration during the transition, and failure to maintain the process after initial implementation. [20–22] Besides, primary care physicians have been tasked as default custodians of a patient’s medication list. [23] However, there are far more opportunities for reconciling medications in routine ambulatory care between primary care, specialists, ambulatory procedures, and retail pharmacies. With the increasing complexity and specialization of medication regimens, it is necessary to expand medication reconciliation at every contact with a clinician. [24]

The collection and availability of vast amounts of detailed patient management data in a variety of information systems, such as electronic prescribing and computerized provider order entry applications, have the potential to facilitate the integration of advanced analytic methodologies, such as Artificial Intelligence (AI) and Operations Research (OR) methods, into supporting the medication reconciliation challenges, thus complementing and enhancing the current process. [4, 25, 26] AI technologies, including data mining, natural language processing, and machine learning, can automate the extraction and analysis of medication data from electronic health records (EHRs). [27] Such automation can streamline the reconciliation process, significantly reducing the time required and minimizing the risk of human error. Additionally, AI systems can continuously learn and adapt, improving their accuracy over time as more data becomes available. [28] Meanwhile, OR can provide robust analytical approaches to optimize the clinical workflow and decision-making processes. This includes the development of algorithms and models that can identify potential discrepancies and suggest corrective actions. [29] Together, AI and OR can transform the way medication reconciliation is conducted, making it more reliable, efficient, scalable, and ultimately enhancing patient safety.

Previous literature reviews have examined the effectiveness of medication reconciliation interventions in clinical settings [30] or the evidence related to screening for medication discrepancies in patient care [31, 32]. However, there has been no explicit review of the problem of improving the medication reconciliation process with AI and OR methods for risk prediction and associated analytics. Other systematic reviews have focused on either specific domains of medication reconciliation, such as error reduction or interventions to improve medication safety among specific disadvantaged groups [33–35], or on specific health outcomes [36, 37], such as length of stay [38], adverse drug events [39], and hospital readmissions [40]. These systematic reviews indicate a dearth of generalizable high-quality evidence on the impact of medication reconciliation interventions using advanced modeling and analytics enabled by AI and OR. Due to the fast-growing use of AI and OR in healthcare and other domains, there is a timely opportunity to understand the factors, data sources, and measures used effectively in AI and OR-enabled medication reconciliation to improve care quality and health outcomes. [26]

This scoping review aims to map the existing research landscape regarding the application of AI and OR in addressing the medication reconciliation problem. We seek to identify and categorize the various methodologies, tools, and frameworks that have been developed and to assess their impact on reducing medication discrepancies and improving patient safety. In this literature review, we explore a wide range of AI and OR applications, from basic rule-based systems to sophisticated machine learning, optimization, and decision-theoretic models. Additionally, we examine case studies to understand the practical implementation and effectiveness of these technologies in real-world healthcare settings.

## Methods

We conducted the scoping review following the guideline Preferred Reporting Items for Systematic Reviews and Meta-Analyses extension for Scoping Reviews (PRISMA-ScR). The PRISMA-ScR checklist is shown in Supporting Information 1. The published literature was searched using strategies created with the help of a university librarian for studies that utilized AI or OR in the medication reconciliation-related context. We used the Sysrev platform to conduct the screening process of the papers. [41]

### Databases and search strategy

We queried PubMed, Web of Science, ACM, INFORMS, IEEE, and CINAHL databases all through December 2023. These databases were selected due to their comprehensive coverage of relevant academic disciplines, including medicine, healthcare, computer science, and operations research. To ensure the inclusiveness and breadth of our search, we designed the search query to be as broad as possible. We adopted concepts related to medication reconciliation, such as medication error, medication discrepancies, adverse drug events, polypharmacy, etc., to cover all pertinent aspects. The search strategies were To find all papers that incorporate AI or OR in medication reconciliation, we carefully established a combination of terminologies to search in the paper title and abstract. The search strategy was as follows:

(ANY medication-reconciliation-related terms in the title or abstract) AND ((ANY AI-related terms in the title or abstract) OR (ANY Operations research-related terms in the title or abstract)), where

- Medication-reconciliation-related terms include: ‘medication reconciliation*’,’medication error*’, medication discrepanc*’, ‘medication accuracy’, ‘adverse drug event*’, ‘polypharmacy’, ‘pharmacovigilance’
- AI-related terms include: ‘artificial intelligence’, ‘machine learning’,’deep learning’, ‘reinforcement learning’, ‘neural network*’, ‘encoder*’, ‘decoder*’, ‘autoencoder*’, ‘generative adversarial network*’, ‘bayes*’, ‘decision tree*’, ‘classifier*’, ‘xgboost’, ‘random forest*’, ‘support vector machine’, ‘bagging’, ‘boosting’, ‘natural language processing’, ‘NLP’, ‘large language model*’, ‘LLM’, ‘generative AI’, ‘GPT’
- OR-related terms include: ‘operation* research’, ‘operation* management’,’integer programming’, ‘linear programming’, ‘mathematical programming’, ‘stochastic programming’, ‘stochastic optimization’, ‘stochastic process’, ‘robust optimization’, ‘Markov’, ‘heuristic*’, ‘metaheuristic*’

A total of 2,000 records were identified by this search strategy. Duplicate records were identified using the automatic deduplicator in SR-Accelerator online review software. [42] We removed 990 duplicates and ended up with 1,100 unique studies for further screening.

### Selection of studies and eligibility criteria

To be eligible for inclusion, published research had to meet all of the following 6 criteria:

1. Peer-reviewed original research with full text published in English.
2. Integrated AI or OR methods into medication reconciliation-related tasks.
3. Examined the impact or performance of integrating AI or OR into medication reconciliation-related tasks.
4. Quantitative analysis.
5. Not limited to the biochemical level of drug reactions.
6. Involved multiple conditions and diseases and not focused on a single medication

Based on these criteria, articles underwent two exclusion steps. Initially, a review of titles and abstracts led to the exclusion of 990 articles. The title and abstract screening excluded systematic reviews/meta-analyses, opinions, commentaries, perspectives, vision articles, guidelines, protocol articles, qualitative studies, exploratory and conceptual studies, and non-English language studies. This was followed by a review of 110 full-text articles, resulting in the exclusion of 46 articles. During the full-text screening, we excluded conference posters. We further checked the relevance of the paper to our topic and only kept studies that focused on leveraging AI/OR in medication reconciliation-related problems. During both exclusion steps, two researchers independently labeled the articles. One researcher was a PhD student majoring in information systems with extensive training in machine learning, optimization, and healthcare, while the other was a graduate student with a medical degree and clinical practice experience. We used the Sysrev online literature review platform to label and record the decisions. [41] Conflicts in labeling were resolved by an experienced researcher specializing in healthcare informatics and analytics. This rigorous process resulted in the inclusion of 64 scientific articles published in the literature. Fig 1 shows the search and screening process of this scoping review. The details of the selected studies are shown in Supporting Information 2.

**Fig 1.**
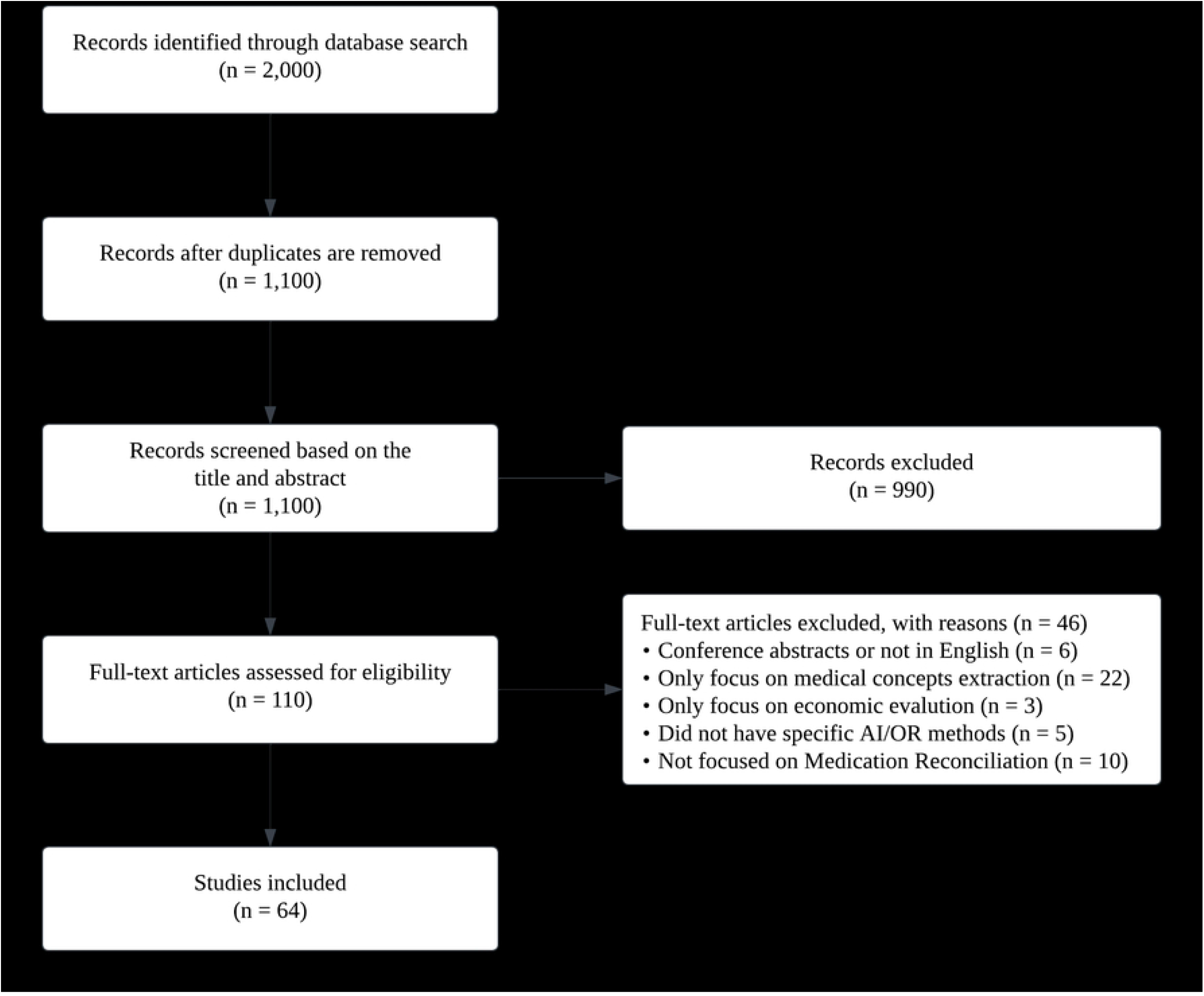
Flowchart of literature search and screening process.

### Data collection and analysis

We collected the 64 articles and extracted different data elements from each, such as year of publication, country of origin, study target, data source, facility and specialty of the data source, data collection start and end time, duration of the data sample, sample size, features included in the data with the total number and feature categories, applied algorithms and their types, outcome measures, outcome performance, and the status of the proposed method. Due to the differences in the included studies and the lack of standardized reporting, we did not conduct a meta-analysis. Instead, we used narrative synthesis to summarize our findings descriptively. To facilitate this process, we utilized ChatGPT [43] to extract and summarize relevant information, followed by meticulous manual review and refinement to ensure analysis accuracy.

## Results

### Overall characteristics of the studies included in this review

We summarize the characteristics of the 64 included studies in Table 1. The temporal distribution of the studies included in this review spans from 2007 to 2023. Early contributions are seen in 2007 with 2 studies, and the number gradually increases to 11 and 13 studies in 2022 and 2023, respectively. We observe that a significant proportion of the studies (45%) published between 2021 and 2023 reflect a recent surge in medication reconciliation-related research activity. This suggests a growing focus and ongoing research in the field of medication errors and adverse events and, thus, medication reconciliation. We also discern a renewed interest in the field with the use of AI and ML methodologies.

**Table 1.** Key characteristics of selected studies

| Characteristics | n | % |
| --- | --- | --- |
| **Publication Year** |  |  |
| 2021-2023 | 29 | 45 |
| 2016-2020 | 20 | 31 |
| 2011-2015 | 12 | 19 |
| 2007-2010 | 3 | 5 |
| **Major Data Sources** |  |  |
| EHRs | 43 | 67 |
| Drug knowledge databases | 20 | 31 |
| ADE reporting systems | 11 | 17 |
| Insurance claims | 4 | 6 |
| **Data Cohort Types** |  |  |
| Outpatient primary care | 6 | 9 |
| Outpatient specialty care | 18 | 28 |
| Surgery/inpatient | 19 | 30 |
| Not specified | 27 | 42 |
| **Data Duration (year)** |  |  |
| (0,1] | 8 | 13 |
| (1,5] | 25 | 39 |
| (5,10] | 9 | 14 |
| (10,20] | 3 | 5 |
| Not specified | 19 | 30 |
| **Application Status** |  |  |
| Prototype | 3 | 5 |
| Experiment | 50 | 78 |
| Deployed | 11 | 17 |

The selected studies primarily relied on Electronic Health Records (EHR) data, with 67% using this source. Drug databases were utilized in almost a third of the studies (31%), showcasing the integration of clinical pharmacology into algorithmic approaches. Adverse Drug Event (ADE) reporting systems featured in 17% of the studies, emphasizing the need to track and mitigate medication-related risks. The data cohort types also varied significantly: 30% of studies focused on surgery/inpatient settings, reflecting the critical need for precise medication management in these complex environments; 28% targeted outpatient specialty care, addressing the diverse medication needs of these patients. However, only 9% of studies covered primary care settings, indicating a lack of sufficient research on medication reconciliation in this fundamental area of healthcare delivery. This distribution demonstrates the varied approaches required for effective medication reconciliation while highlighting the need for more research in primary care.

In terms of data sample sizes, the studies also demonstrated considerable variation, shown in Fig 2. 14 studies employed sample sizes in the range of 100 to 10,000 patients, reflecting a focus on moderately sized datasets. Smaller datasets with 0 to 100 patients were used in 5 studies, likely for initial exploratory research. Larger datasets, containing 10,000 to 1,000,000 patients, were leveraged in 12 studies, while 8 studies utilized very large datasets ranging from 1,000,000 to 10,000,000 patients. This diversity in data sample sizes highlights the increasing capacity for large-scale data analytics in medication reconciliation-related quantitative research, enhancing the robustness of patient medication management insights.

**Fig 2.**
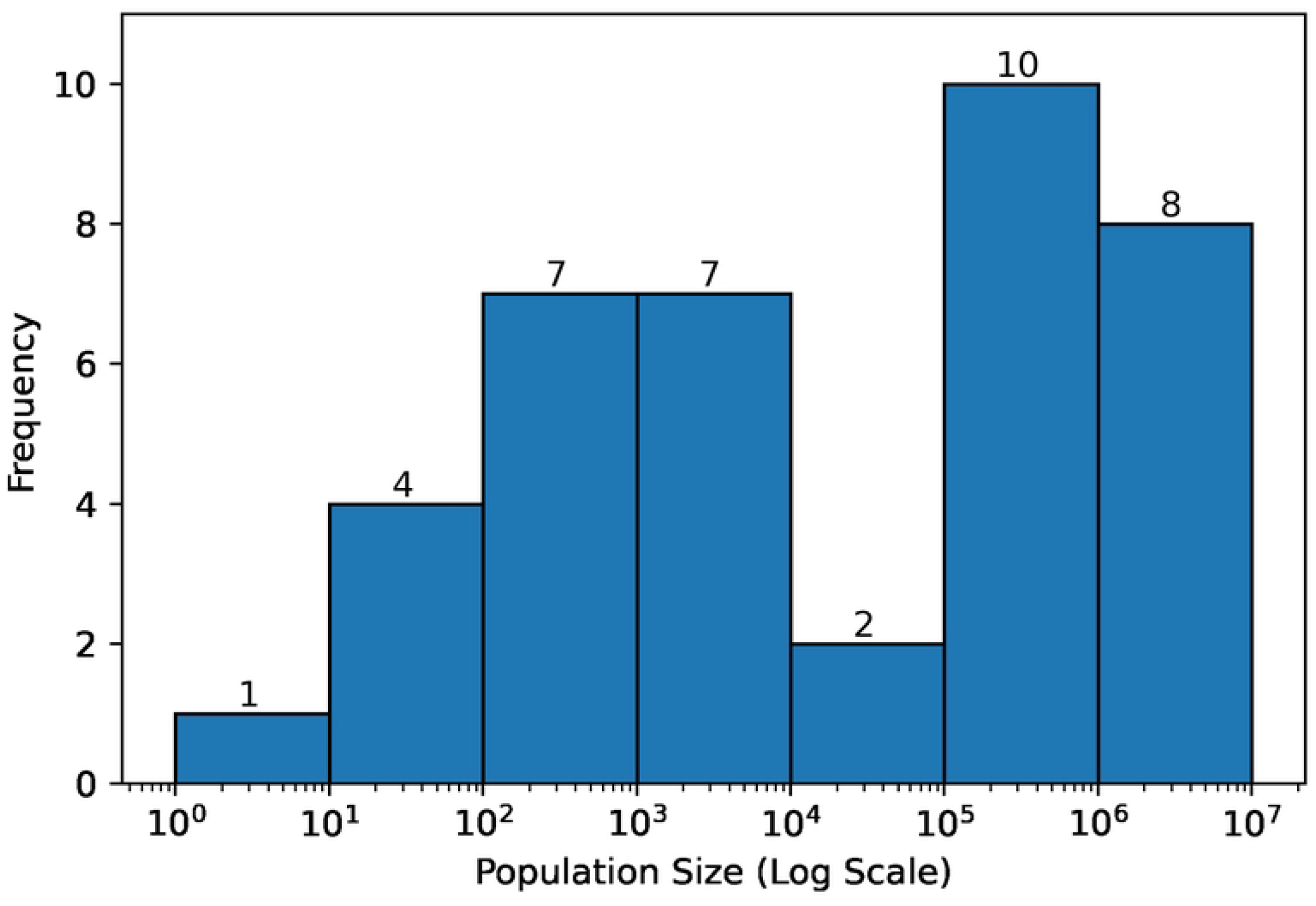
Histogram of patient population size in the included studies (log scale)

Regarding data duration, several studies collected data for 1 to 5 years (39%), while only 5% of studies collected data for more than 10 years. The most commonly encountered study duration was two years, represented by 9 studies, suggesting a preference for medium-term studies on this research topic. Although longer-term studies likely provide more comprehensive insights, they are fewer in number due to the extended time commitments and resources required.

Most studies were in the experimental phase (80%), which focuses on validation and testing within the context of medication reconciliation or related topics using AI/OR methods. Fewer studies were in the prototype stage (5%) or had reached deployed status (16%), indicating a continuing need for medication reconciliation research to be deployed and evaluated in real-world clinical settings.

### Geographic regions of the selected research

The selected set of papers analyzed in this review reflects a broad international interest in the subject matter. The United States leads with the highest number of studies, contributing 17 papers, closely followed by Sweden with 14 studies. France and the Greater China region also contribute a number of papers, with 6 and 5 studies, respectively. Denmark follows with 3 studies, while several other countries, including Australia, Canada, India, Iran, Japan, and Turkey, each contribute 2 studies. The country of origin was not clearly specified in 5 studies. At the continent level, as shown in Fig 3, Europe has the highest number of papers related to medication reconciliation topics using AI and OR methods, followed by North America, Asia, and Australia, in that order. No related papers were found from Africa and South America.

**Fig 3.**
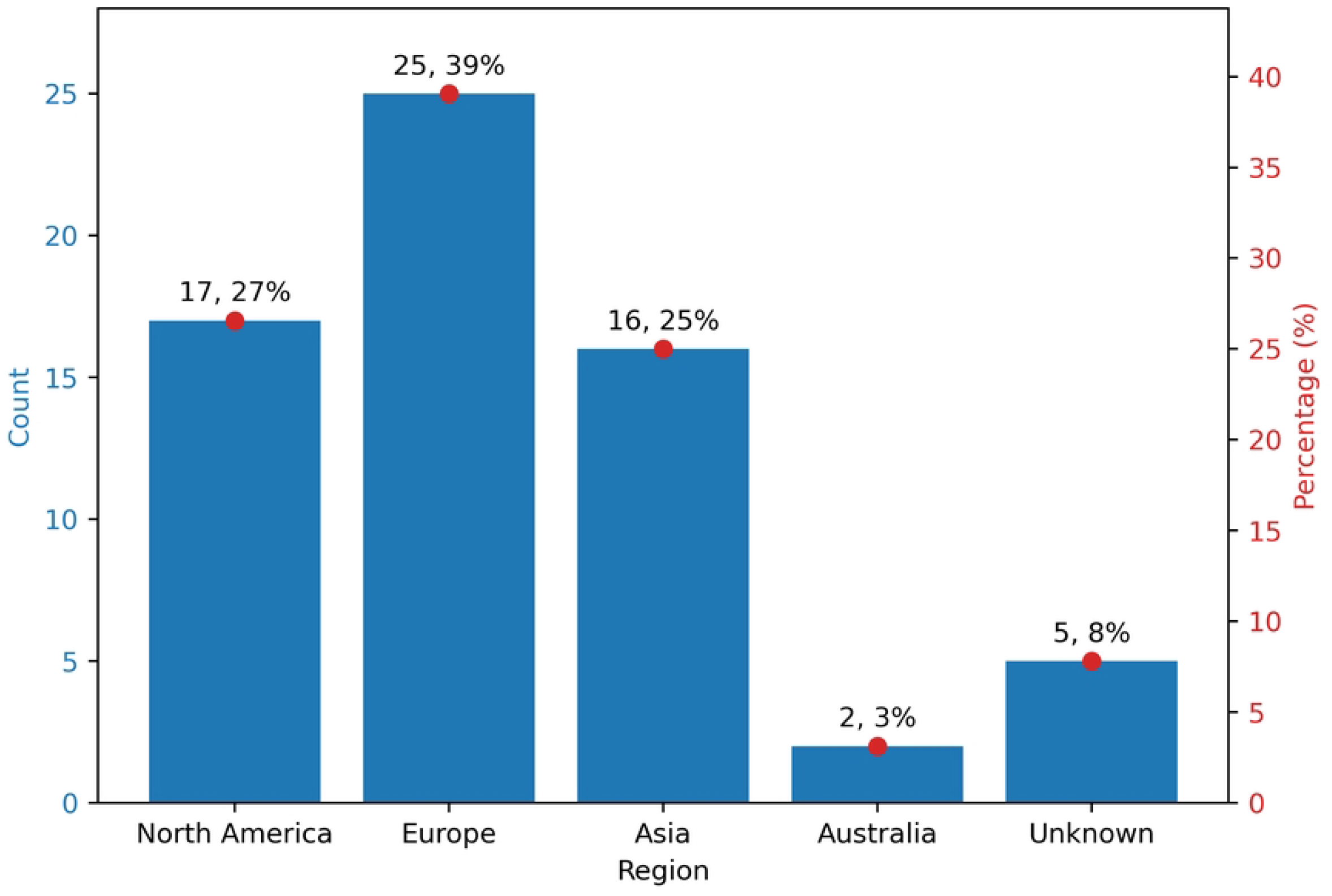
Distribution of the selected studies across different continents.

We note that only 3 papers (5%) utilize data from different geographic regions, indicating a lack of geographical diversity for medication reconciliation studies. [44–46] This limitation also presents challenges to the generalizability and applicability of the proposed algorithms and models in the literature. When algorithms are developed and validated using data from a single region, they may not perform as effectively in different regional healthcare settings with varying patient populations, healthcare practices, and data characteristics. This geographical constraint underscores the need for more collaborative, multi-regional studies to enhance the robustness and versatility of medication reconciliation solutions. Expanding research to include diverse data sources from multiple regions will help ensure that the developed models are more universally applicable, ultimately leading to improved patient safety and medication management in varied healthcare environments.

### Topics and analytic approaches used in the selected papers

The research topics of the selected papers reveal several key priorities. ADE Detection, at 27%, is the largest area of focus, highlighting its critical role in ensuring medication reconciliation by identifying recorded adverse events in the medical history. [45–61] Medication error detection follows closely at 23%, which emphasizes identifying and correcting errors in medication administration. [4, 44, 62–73] ADE prediction, representing 22%, reflects proactive efforts to foresee and prevent adverse events before they occur. [74–87] Polypharmacy management, at 9%, addresses the complexities involved in managing patients who are prescribed multiple medications, which is crucial for reducing drug interactions and improving outcomes. [88–93] Studies directly addressing medication reconciliation account for only 8%, indicating that this critical area remains understudied. [15, 94–97] This limited focus suggests a need for more comprehensive research to ensure accurate, comprehensive, and consistent medication records across different healthcare settings for every patient, which is essential for minimizing discrepancies and enhancing patient safety. Medication review, constituting 5%, focuses on the regular assessment of patients’ medication regimens to ensure their appropriateness and effectiveness. [98–100] Finally, Drug dosage adjustment [101, 102] and medication recommendation [103, 104] each make up 3% of the selected studies, highlighting the need for tailored dosage modifications and personalized medication advice to optimize treatment. The topic distribution of the selection papers is summarized in Figure 4.

**Fig 4.**
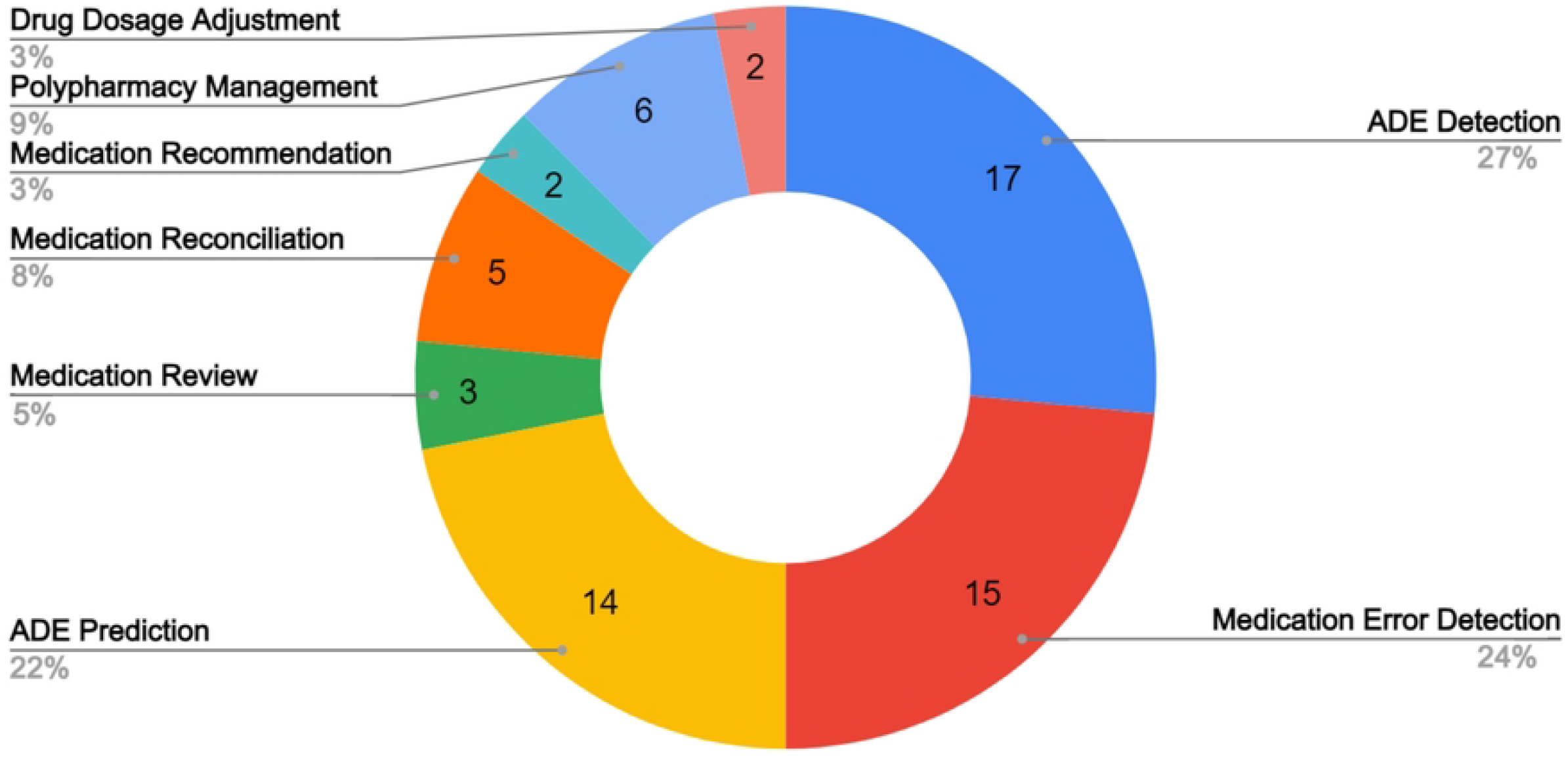
Pie chart of the topics of the selected paper.

We also summarize the analytic approaches used in the selected literature in Fig 5. The methods have been categorized into the following groups: tree-based ML models, regression-based ML models, support vector machines, Natural Language Processing (NLP) algorithms, non-NLP deep learning (DL) models, and decision-theoretic methods. The graph shows that the most frequently used methods are tree-based approaches, which are featured in 33 studies. NLP techniques are used in 27 studies, highlighting the increasing importance of text and language data in EHRs and clinical notes.

**Fig 5.**
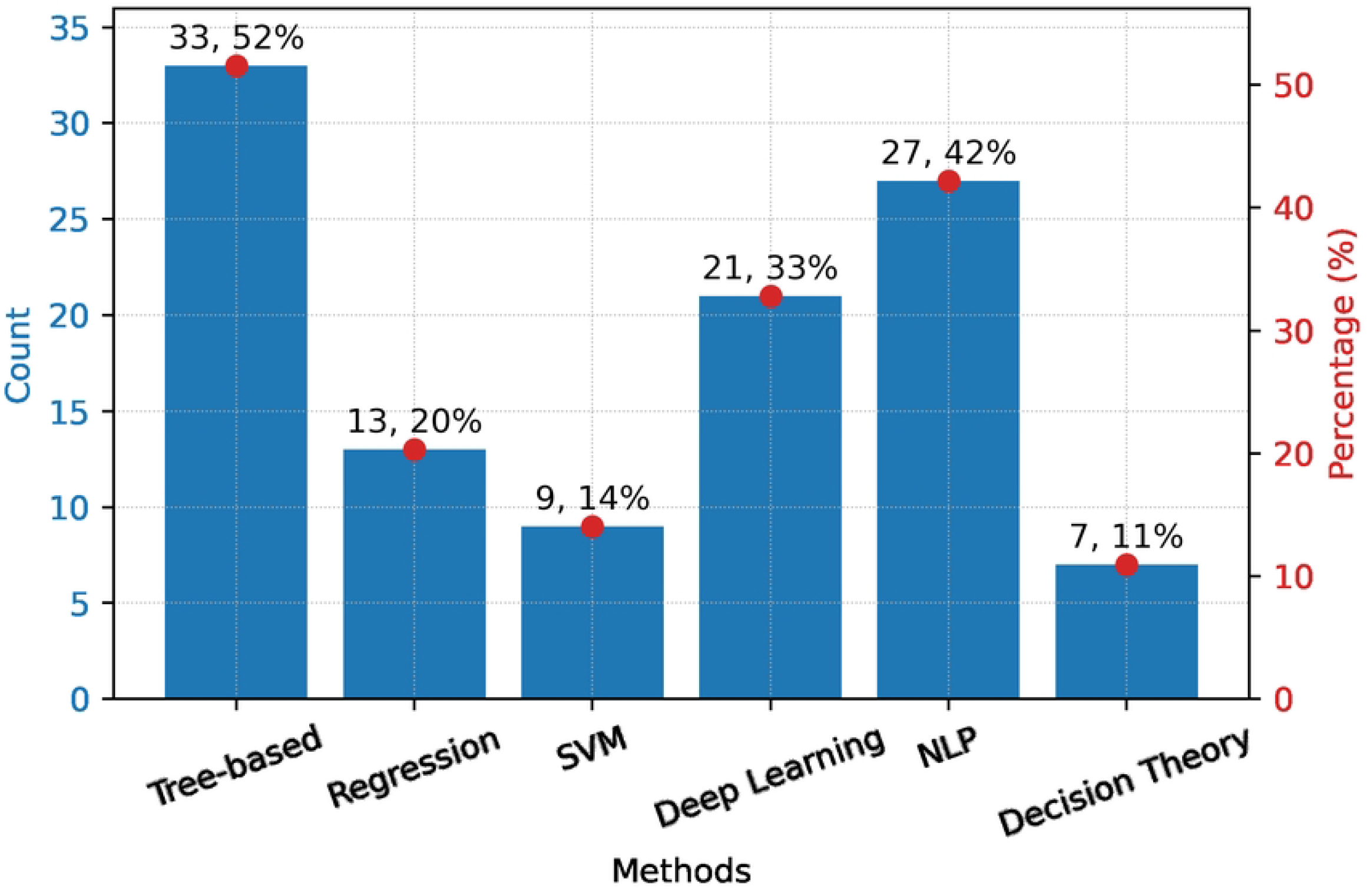
Algorithms used in the selected literature.

However, as of the end of 2023, we identified only one selected paper that applies Large Language Models (LLM) for medication error classification, highlighting a significant research opportunity in medication reconciliation with LLMs. [63] Non-NLP DL methods are used in 21 studies. Regression-based methods are used in 13 studies, and SVM is featured in 9 studies. Decision Theory approaches are employed in 7 studies, illustrating the limited use of formal decision-making frameworks in medication reconciliation research.

In summary, our review confirms the diverse methodologies and topics explored in existing studies. However, there remains a significant need for the integration of advanced algorithms into clinical information systems. Additionally, the generalizability of these algorithms is limited due to the reliance on data from single or dual EHR systems within specific geographic regions, posing challenges for broader application across different healthcare facilities.

## Discussion

### Current efforts for medication reconciliation with AI and OR

AI-based approaches are revolutionizing the field of medication reconciliation, with their great potential to enhance precision, efficiency, and accuracy compared to traditional manual methods. [4, 48, 64, 88, 94, 105] Historically, the medication reconciliation process has been labor-intensive and prone to human error due to its reliance on manual checks. [66, 106, 107] The integration of AI technologies is transforming this landscape by introducing more reliable, consistent, and streamlined workflows, significantly improving the overall effectiveness of medication reconciliation and thereby bolstering patient safety.

A key application of AI in medication reconciliation is through NLP. Given that EHRs are rich in patient data, much of which is stored in unstructured formats like physician notes, discharge summaries, and clinical narratives, traditionally, extracting relevant medication information from these documents was a time-consuming and error-prone task. [49, 61, 98] NLP algorithms can transform this aspect of medication reconciliation by automating the interpretation and extraction of essential information from unstructured text, such as drug names, dosages, frequencies, administration routes, conditions, and adverse events. [94] This structured extracted data can then be seamlessly integrated with other EHR components, enabling the identification of discrepancies, such as mismatches between physician notes and formal prescription orders, and allowing for immediate corrective actions. [76, 88] The automation provided by NLP not only reduces the workload on healthcare providers but also significantly enhances the accuracy and speed of the medication reconciliation process. [15, 63] Among various NLP algorithms, Conditional Random Fields (CRFs) historically have been a dominant approach for detecting ADE and medication discrepancies within clinical texts through named entity recognition and relation extraction in the medical domain. [53, 94] Although laying the groundwork for sequence labeling, and continuing to be used as a valuable tool, CRFs have been largely supplanted by or combined with more advanced models, such as self-attention models, that offer greater accuracy and contextual understanding. Recent advancements have shifted focus toward deep learning models, particularly those based on transformer architectures like BERT (Bidirectional Encoder Representations from Transformers). [52–54, 63, 98, 108] With deep contextual understanding, these advanced NLP models can help discern that certain medications should not be co-prescribed or recognize potential omissions in a medication list based on a patient’s medical history. They can also handle complex cases involving multiple medications and conditions with greater accuracy, reducing the risk of oversight and ensuring that patients receive the correct medications.

Beyond NLP-related models, machine learning approaches such as regression models and tree-based techniques are integral to predictive analytics and decision-making in medication reconciliation. [48, 65, 66, 84] Regression models, traditionally used for predicting continuous outcomes, can also be employed for classification, helping to identify the likelihood of various outcomes. In the context of medication reconciliation, regression models can predict the probability of experiencing potential adverse drug events and quantify the impact of various patient factors, such as age, weight, and existing health conditions, allowing for more nuanced and personalized predictions. [75, 99] Tree-based models, including ID3 decision trees, random forests, and gradient-boosted trees, excel in making predictions through data-driven splits, effectively handling classification tasks for medication reconciliation. [50, 89, 109] These models are particularly valuable for modeling relationships between a dependent variable and multiple independent variables, providing insights into the strength and nature of these relationships, which are crucial in analyzing large datasets that include patient demographics, comorbidities, and historical medication usage. [62, 64, 88] By uncovering patterns and correlations that may not be immediately obvious to clinicians, these models can forecast potential adverse events, flag possible drug interactions, and recommend appropriate medication adjustments. For instance, a regression analysis might reveal a higher risk of adverse reactions in a particular patient group, guiding safer prescribing practices. [84, 102] Moreover, both tree-based and regression models are often preferred for their interpretability compared to more complex neural networks. [110] Their transparent decision-making process allows healthcare providers to understand and trust the rationale behind AI-generated recommendations. [111] Incorporating these models into the medication reconciliation process may enable healthcare providers to anticipate risks, customize treatment plans, and suggest safer alternatives, ultimately improving patient safety and treatment outcomes.

As for the selected decision-theoretic papers, mostly of them are rule-based decision support models, which do not cover common OR techniques, such as mathematical optimization and simulation. [92, 99] Although the literature on the decision-theoretic OR models in medication reconciliation is limited, OR has the potential to significantly impact medication reconciliation by providing data-driven, optimized approaches to streamline the process, prioritize high-risk patients for manual review, optimize the allocation of healthcare resources, and reduce the cognitive load on clinicians. [4, 26, 105, 112] While AI models can predict high-risk scenarios, OR methods could optimize the response by designing efficient workflows to mitigate those risks. [113] This could lead to faster detection of medication errors, minimizing adverse drug events, and improving patient safety. [114]

### Technology-enabled medication reconciliation-related solutions

The limited real-world deployment and application of models developed in the medication reconciliation literature reveals an opportunity to translate theoretical insights into clinical improvements. The literature indicates that Clinical Decision Support Systems (CDSS) applied in medication reconciliation can primarily be categorized into two areas: risk alerts and drug recommendations. In risk alert applications, typical scenarios include polypharmacy management and error prevention. As for polypharmacy, CDSS tools alert clinicians to potential drug-drug interactions or duplications, thereby reducing side effects and improving patient outcomes [80, 88, 92]. Error prevention tools, such as MedAware, focus on identifying outliers in prescriptions based on patient-specific clinical data, preventing medication errors, and enhancing patient safety [69, 70, 72]. Besides, AI-powered prescription alert systems are also shown to significantly reduce irrelevant alerts compared to rule-based Computerized Physician Order Entry (CPOE) systems, streamlining the clinical decision-making process [69, 92]. In drug recommendation applications, e-prescribing systems integrate recommendations to reconcile potential discrepancies and flag adverse interactions, especially in long-term care [104]. Advanced AI-based tools further improve accuracy by offering real-time predictions that can account for allergy information and drug incompatibilities [15, 68, 97]. However, integrating these tools into existing clinical workflows remains challenging, limiting broader applicability and clinician acceptance. Concerns around the transparency of AI algorithms and the risk of alert fatigue also complicate their implementation.

In addition to literature-based models, we also found some commercial medication reconciliation tools that have been integrated into healthcare systems, enhancing patient safety by reducing manual data entry and providing a consistent view of patient medication histories. Tools like DrFirst’s Rcopia [115] and Cureatr’s Medication Management Software [116] leverage AI to enhance data usability and alert capabilities. Rcopia uses AI to interpret free-text prescriptions, while Cureatr provides real-time alerts for drug interactions. However, their effectiveness depends on data quality, and complex clinical judgments may still require clinician oversight. Other solutions, such as Avicenna Medical Systems’ MedRec [117] and Health Care Systems’ HCS Medication Reconciliation [118], focus on data integration and workflow automation. MedRec, used extensively within Veterans Affairs hospitals, consolidates data across sources but lacks predictive capabilities, limiting proactive discrepancy identification. HCS’s tool emphasizes efficiency by automating medication history retrieval using static algorithms, but it may lack flexibility in adapting to unique clinical cases. Tools such as MEDITECH’s Expanse Pharmacy [119] and WellSky Medication Management [120] emphasize workflow customization and support, though extensive training is often necessary for full functionality. Systems like Better Meds [121] and First Databank’s MedKnowledge [122] offer comprehensive data integration and drug information, but challenges with disparate technologies can affect their interoperability in some settings. Platforms such as RXNT [123] offer extensive functionalities and support for medication management. However, limitations in interoperability with other electronic health systems can hinder their seamless integration and usability in diverse clinical environments.

Overall, both the selected literature and commercial tools underscore the importance of tech-enabled solutions in improving medication safety and prescribing efficiency.

Despite their benefits, challenges such as EHR interoperability and data quality remain, necessitating ongoing validation to ensure responsiveness to evolving clinical needs.

Continuous real-world application is essential to bridge the gap between theoretical models and practical implementation for improving the medication reconciliation process.

### Challenges in medication reconciliation with advanced AI and OR approaches

Despite significant advancements in both AI and OR, their development and integration in the medication reconciliation pipeline remain limited. The challenges result from the complex and fragmented nature of healthcare systems. One major barrier to effectively integrating both AI and OR in medication reconciliation is data quality. [124] Incomplete, inaccurate, or biased EHR data can reduce the accuracy of ML models, which may lead to false positives or negatives when identifying medication discrepancies. [125] Similarly, with inaccurate input data in the OR models, the optimization programs will yield a suboptimal decision for the real-world system. [126] This highlights the need for robust data governance and continuous improvements in healthcare data practices to ensure that AI and OR can work together effectively.

Besides, integrating AI and OR systems into the existing EHR workflows faces transparency and interoperability issues. The ‘black-box’ nature of many AI models raises concerns about reliability, which is critical for clinical decision-making. [127] The lack of clear model explanations can reduce trust in these systems, reinforcing the need for explainable AI methods, alongside OR-driven optimization, to enhance their clinical utility.

In summary, efforts to integrate AI and OR are still in the early stages, which indicates a promising direction for hybrid models that combine AI to predict discrepancies and OR to design real-time interventions. However, more research is needed to overcome challenges like data interoperability, system integration, and workflow redesign. These technical and logistical barriers must be addressed to fully unlock the potential of AI and OR for medication reconciliation in healthcare.

### Clinician perspectives

AI-powered tools can automate data extraction from EHRs and reduce administrative tasks for clinicians, allowing them to focus more on clinical decisions. [49, 74, 91] Although several of the published studies focus on medication error management, a majority of them concentrate on single drugs or a narrow panel prescribed within a specific specialty. [128, 129] Ideally, reconciliation should account for all medications a patient may be taking, including over-the-counter (OTC) and also unprescribed substances. [130] A clinical decision support system that could predict what non-prescribed medications a patient is likely to take would be highly beneficial, which can enhance the comprehensiveness of medication reconciliation and reduce the risk of missing important interactions. [131]

Almost 72% of the studies reviewed centered on ADE detection/prediction or medical error detection, which is important but represents only one step in the overall medication reconciliation process. A very small number of studies explored the complete process, highlighting a gap in the research. [15, 94–97] Additionally, the challenge of alert fatigue, a common issue when introducing technological solutions to clinical workflows, was not adequately addressed in most studies. [132] This is a crucial consideration, as an effective medication reconciliation solution must balance comprehensive alerts with minimizing unnecessary interruptions to clinical care.

Moreover, the review identifies a gap in the use of OR methodologies that can optimize clinical workflows and reduce the burden on clinicians. Decision-theoretic models and other OR techniques remain underutilized. [26] More research is necessary, not just on developing sophisticated algorithms, but on how they can be seamlessly integrated into everyday clinical workflows, making medication reconciliation solutions practical for adoption and daily use. [133, 134]

### Limitations and Future Work

There are limitations to our review. Although we searched for articles across 5 large databases, we only considered the published literature and, therefore, may have failed to capture unpublished studies that might also contain relevant content. As considerable studies on medication reconciliation and AI/OR algorithms are disseminated through conferences, posters, and other non-traditional means, which are often not indexed in standard bibliographic databases, our focus on bibliographic databases may have introduced selection bias by excluding potentially valuable insights found in these alternative sources.

Furthermore, the majority of the identified publications evaluated algorithms at the single-drug or chemical level and were consequently excluded from our review. While we believe that this exclusion does not fundamentally alter the overall content or methodologies of the medication reconciliation efforts examined, it may impact the perceived comprehensiveness of our findings. This filtering process, while emphasizing studies involving multiple medications and symptoms, may inadvertently overlook valuable insights specific to single-drug evaluations, which could contribute to a more nuanced understanding of AI and OR applications in medication reconciliation. Besides, the executed search strategy required iteration due to the evolving terminology in AI/OR-driven biomedicine research, necessitating the inclusion of new terms and definitions over time.

From the analysis perspective, we recommend that future research delve deeper into advanced AI topics. Our review primarily focused on traditional machine learning categories, such as tree-based, regression-based, NLP-based, and deep learning models. However, we did not place significant emphasis on categories such as generative AI and large language models (LLMs) due to the limited number of papers available on this topic at the time of our review, even though we included LLM-related keywords in our search strategy. This relative scarcity of studies in this area suggests an opportunity for further exploration as LLMs continue to evolve and gain traction in healthcare applications, particularly in medication reconciliation.

## Conclusion

This review contributes to the existing body of knowledge by examining the current usage of AI and OR approaches for improving medication reconciliation and clarifying how AI and OR can be effectively utilized to streamline medication reconciliation processes in the future. We highlight the necessity for continuous research and development, urging healthcare organizations to embrace these innovative solutions.

Addressing the barriers to effective implementation and identifying best practices are critical steps toward enabling the widespread integration of AI and OR in medication reconciliation systems. Despite these advancements, our findings reveal a substantial disparity between the theoretical accuracy of algorithms and their practical application at the healthcare decision-making level. Few studies have moved beyond theoretical frameworks to implement these applications in real clinical settings. This gap underscores the need for further research that not only measures but also enhances the integration of these algorithms into everyday healthcare workflows. By addressing this gap, we can ensure that AI and OR technologies do not just exist in theory but are actively improving patient care and safety.

In conclusion, this scoping review outlines the potential and current state of AI and OR in medication reconciliation. The integration of these technologies represents a significant advancement in healthcare management, offering a pathway to reduce ADEs and improve patient outcomes through automation of risk assessment and optimization of real-time decision making capability. The review highlights the benefits, challenges, and future directions of AI and OR in this field, providing a comprehensive overview for researchers, practitioners, and policymakers. Continued collaboration and research are essential to address the remaining challenges and fully realize the potential of these technologies, ultimately enhancing patient safety and healthcare quality.

## Data Availability

All relevant data are within the manuscript and its Supporting Information files.

## Author Contributions

RP conceptualized the idea for this study. XY and RP designed it and framed reference search strategies. XY searched the databases and conducted record deduplication. XY and AR performed screening, study selection, data extraction, and data analysis, with extensive input and feedback from RP. XY contributed to the original draft, with input on the clinical perspectives from AR. RP performed review and editing. RP supervised the study. All authors approved the final draft for submission.

## Acknowledgements

We thank Sarah Young at Carnegie Mellon University Libraries for assistance with the literature search resources.

## References

1. Kuperman GJ, Bobb A, Payne TH, Avery AJ, Gandhi TK, Burns G, et al. Medication-related clinical decision support in computerized provider order entry systems: a review. Journal of the American Medical Informatics Association. 2007;14(1):29–40.

2. Bennett S. WHO launches global effort to halve medication-related errors in 5 years. World Health Organization, Department of Communications Geneva: WHO Department of Communications. 2017;.

3. Masnoon N, Shakib S, Kalisch-Ellett L, Caughey GE. What is polypharmacy? A systematic review of definitions. BMC geriatrics. 2017;17:1–10.

4. Hasan S, Duncan GT, Neill DB, Padman R. Automatic detection of omissions in medication lists. Journal of the American Medical Informatics Association. 2011;18(4):449–458.

5. Sternberg SA, Guy-Alfandary S, Rochon PA. Prescribing cascades in older adults. CMAJ. 2021;193(6):E215–E215.

6. Reidenberg MM. Diagnosis drift and its contribution to polypharmacy. Clinical Pharmacology & Therapeutics. 2018;103(4):556–557.

7. Bates DW. Preventing medication errors: a summary. American Journal of Health-System Pharmacy. 2007;64(14 Supplement 9):S3–S9.

8. Longhurst CA, Parast L, Sandborg CI, Widen E, Sullivan J, Hahn JS, et al. Decrease in hospital-wide mortality rate after implementation of a commercially sold computerized physician order entry system. Pediatrics. 2010;126(1):14–21.

9. Sittig DF, Belmont E, Singh H. Improving the safety of health information technology requires shared responsibility: It is time we all step up. In: Healthcare. vol. 6. Elsevier; 2018. p. 7–12.

10. Alert SE. Using medication reconciliation to prevent errors. Journal on Quality and Patient Safety. 2006;32(4):230–2.

11. CMS. Eligible professional meaningful Use menu set measures: Measure 6 of 9; 2014. https://www.cms.gov/Regulations-and-Guidance/Legislation/EHRIncentivePrograms/downloads/7_Medication_Reconciliation.pdf.

12. Almanasreh E, Moles R, Chen TF. The medication reconciliation process and classification of discrepancies: a systematic review. British journal of clinical pharmacology. 2016;82(3):645–658.

13. Kwan JL, Lo L, Sampson M, Shojania KG. Medication reconciliation during transitions of care as a patient safety strategy: a systematic review. Annals of internal medicine. 2013;158(5 Part 2):397–403.

14. Mueller SK, Sponsler KC, Kripalani S, Schnipper JL. Hospital-based medication reconciliation practices: a systematic review. Archives of internal medicine. 2012;172(14):1057–1069.

15. Long J, Yuan MJ, Poonawala R, et al. An observational study to evaluate the usability and intent to adopt an artificial intelligence–powered medication reconciliation tool. Interactive journal of medical research. 2016;5(2):e5462.

16. Santell JP. Reconciliation failures lead to medication errors. Joint Commission journal on quality and patient safety. 2006;32(4):225–229.

17. Slight SP, Tolley CL, Bates DW, Fraser R, Bigirumurame T, Kasim A, et al. Medication errors and adverse drug events in a UK hospital during the optimisation of electronic prescriptions: a prospective observational study. The Lancet Digital Health. 2019;1(8):e403–e412.

18. Barnsteiner JH. Medication Reconciliation: Transfer of medication information across settings—keeping it free from error. AJN The American Journal of Nursing. 2005;105(3):31–36.

19. Rungvivatjarus T, Kuelbs CL, Miller L, Perham J, Sanderson K, Billman G, et al. Medication reconciliation improvement utilizing process redesign and clinical decision support. The Joint Commission Journal on Quality and Patient Safety. 2020;46(1):27–36.

20. Glintborg B, Andersen SE, Dalhoff K. Insufficient communication about medication use at the interface between hospital and primary care. BMJ Quality & Safety. 2007;16(1):34–39.

21. van Sluisveld N, Zegers M, Natsch S, Wollersheim H. Medication reconciliation at hospital admission and discharge: insufficient knowledge, unclear task reallocation and lack of collaboration as major barriers to medication safety. BMC health services research. 2012;12:1–12.

22. Salanitro AH, Osborn CY, Schnipper JL, Roumie CL, Labonville S, Johnson DC, et al. Effect of patient-and medication-related factors on inpatient medication reconciliation errors. Journal of general internal medicine. 2012;27:924–932.

23. Kashyap N, Jeffery S, Agresta T. From MedWreck to MedRec: A Call to Action to Improve Medication Reconciliation. Applied Clinical Informatics. 2024;15(02):230–233.

24. Varkey P, Cunningham J, Bisping S. Improving medication reconciliation in the outpatient setting. The Joint Commission Journal on Quality and Patient Safety. 2007;33(5):286–292.

25. Choudhury A, Asan O, et al. Role of artificial intelligence in patient safety outcomes: systematic literature review. JMIR medical informatics. 2020;8(7):e18599.

26. Kruik-Kollöffel W, Moltman G, Wu M, Braaksma A, Karapinar F, Boucherie R. Optimisation of medication reconciliation using queueing theory: a computer experiment. International Journal of Clinical Pharmacy. 2024; p. 1–8.

27. Zhu R, Tu X, Huang J. Using deep learning based natural language processing techniques for clinical decision-making with EHRs. Deep learning techniques for biomedical and health informatics. 2020; p. 257–295.

28. Alowais SA, Alghamdi SS, Alsuhebany N, Alqahtani T, Alshaya AI, Almohareb SN, et al. Revolutionizing healthcare: the role of artificial intelligence in clinical practice. BMC medical education. 2023;23(1):689.

29. Guha S, Kumar S. Emergence of big data research in operations management, information systems, and healthcare: Past contributions and future roadmap. Production and Operations Management. 2018;27(9):1724–1735.

30. Anderson LJ, Schnipper JL, Nuckols TK, Shane R, Le MM, Robbins K, et al. Effect of medication reconciliation interventions on outcomes: a systematic overview of systematic reviews. American Journal of Health-System Pharmacy. 2019;76(24):2028–2040.

31. Kee KW, Char CWT, Yip AYF. A review on interventions to reduce medication discrepancies or errors in primary or ambulatory care setting during care transition from hospital to primary care. Journal of family medicine and primary care. 2018;7(3):501–506.

32. Alqenae FA, Steinke D, Keers RN. Prevalence and nature of medication errors and medication-related harm following discharge from hospital to community settings: a systematic review. Drug safety. 2020;43:517–537.

33. Chauhan A, Walton M, Manias E, Walpola RL, Seale H, Latanik M, et al. The safety of health care for ethnic minority patients: a systematic review. International journal for equity in health. 2020;19:1–25.

34. Cheraghi-Sohi S, Panagioti M, Daker-White G, Giles S, Riste L, Kirk S, et al. Patient safety in marginalised groups: a narrative scoping review. International journal for equity in health. 2020;19:1–26.

35. Naseralallah LM, Hussain TA, Jaam M, Pawluk SA. Impact of pharmacist interventions on medication errors in hospitalized pediatric patients: a systematic review and meta-analysis. International journal of clinical pharmacy. 2020;42(4):979–994.

36. Guisado-Gil AB, Mejías-Trueba M, Alfaro-Lara ER, Sánchez-Hidalgo M, Ramírez-Duque N, Santos-Rubio MD. Impact of medication reconciliation on health outcomes: An overview of systematic reviews. Research in Social and Administrative Pharmacy. 2020;16(8):995–1002.

37. Lehnbom EC, Stewart MJ, Manias E, Westbrook JI. Impact of medication reconciliation and review on clinical outcomes. Annals of Pharmacotherapy. 2014;48(10):1298–1312.

38. Siddique SM, Tipton K, Leas B, Greysen SR, Mull NK, Lane-Fall M, et al. Interventions to reduce hospital length of stay in high-risk populations: a systematic review. JAMA network open. 2021;4(9):e2125846–e2125846.

39. Hailu BY, Berhe DF, Gudina EK, Gidey K, Getachew M. Drug related problems in admitted geriatric patients: the impact of clinical pharmacist interventions. BMC geriatrics. 2020;20:1–8.

40. Renaudin P, Boyer L, Esteve MA, Bertault-Peres P, Auquier P, Honore S. Do pharmacist-led medication reviews in hospitals help reduce hospital readmissions? A systematic review and meta-analysis. British journal of clinical pharmacology. 2016;82(6):1660–1673.

41. Bozada Jr T, Borden J, Workman J, Del Cid M, Malinowski J, Luechtefeld T. Sysrev: a FAIR platform for data curation and systematic evidence review. Frontiers in Artificial Intelligence. 2021;4:685298.

42. Forbes C, Greenwood H, Carter M, Clark J. Automation of duplicate record detection for systematic reviews: Deduplicator. Systematic reviews. 2024;13(1):206.

43. OpenAI. GPT-4 is OpenAI’s most advanced system, producing safer and more useful responses; 2023. Available from: https://openai.com/index/gpt-4/.

44. Chin YPH, Song W, Lien CE, Yoon CH, Wang WC, Liu J, et al. Assessing the international transferability of a machine learning model for detecting medication error in the general internal medicine clinic: multicenter preliminary validation study. JMIR Medical Informatics. 2021;9(1):e23454.

45. Chazard E, Ficheur G, Bernonville S, Luyckx M, Beuscart R. Data mining to generate adverse drug events detection rules. IEEE Transactions on Information Technology in Biomedicine. 2011;15(6):823–830.

46. Chazard E, Preda C, Merlin B, Ficheur G, Beuscart R. Data-mining-based detection of adverse drug events. In: Medical Informatics in a United and Healthy Europe. IOS Press; 2009. p. 552–556.

47. Bagattini F, Karlsson I, Rebane J, Papapetrou P. A classification framework for exploiting sparse multi-variate temporal features with application to adverse drug event detection in medical records. BMC medical informatics and decision making. 2019;19:1–20.

48. Wang G, Jung K, Winnenburg R, Shah NH. A method for systematic discovery of adverse drug events from clinical notes. Journal of the American Medical Informatics Association. 2015;22(6):1196–1204.

49. Geva A, Stedman JP, Manzi SF, Lin C, Savova GK, Avillach P, et al. Adverse drug event presentation and tracking (ADEPT): semiautomated, high throughput pharmacovigilance using real-world data. JAMIA open. 2020;3(3):413–421.

50. Zhao J, Henriksson A, Boström H. Cascading adverse drug event detection in electronic health records. In: 2015 IEEE International Conference on Data Science and Advanced Analytics (DSAA). IEEE; 2015. p. 1–8.

51. Zhao J, Henriksson A, Asker L, Boström H. Detecting adverse drug events with multiple representations of clinical measurements. In: 2014 IEEE International Conference on Bioinformatics and Biomedicine (BIBM). IEEE; 2014. p. 536–543.

52. McMaster C, Chan J, Liew DF, Su E, Frauman AG, Chapman WW, et al. Developing a deep learning natural language processing algorithm for automated reporting of adverse drug reactions. Journal of biomedical informatics. 2023;137:104265.

53. Feng ZY, Wu XH, Ma JL, Li M, He GF, Cao DS, et al. DKADE: a novel framework based on deep learning and knowledge graph for identifying adverse drug events and related medications. Briefings in Bioinformatics. 2023;24(4):bbad228.

54. Zitu MM, Zhang S, Owen DH, Chiang C, Li L. Generalizability of machine learning methods in detecting adverse drug events from clinical narratives in electronic medical records. Frontiers in Pharmacology. 2023;14:1218679.

55. Henelius A, Puolamäki K, Karlsson I, Zhao J, Asker L, Boström H, et al. Goldeneye++: A closer look into the black box. In: Statistical Learning and Data Sciences: Third International Symposium, SLDS 2015, Egham, UK, April 20-23, 2015, Proceedings 3. Springer; 2015. p. 96–105.

56. Zhao J, Henriksson A, Kvist M, Asker L, Boström H. Handling temporality of clinical events for drug safety surveillance. In: AMIA Annual Symposium Proceedings. vol. 2015. American Medical Informatics Association; 2015. p. 1371.

57. Iqbal E, Mallah R, Jackson RG, Ball M, Ibrahim ZM, Broadbent M, et al. Identification of adverse drug events from free text electronic patient records and information in a large mental health case register. PloS one. 2015;10(8):e0134208.

58. Wasylewicz A, van de Burgt B, Weterings A, Jessurun N, Korsten E, Egberts T, et al. Identifying adverse drug reactions from free-text electronic hospital health record notes. British Journal of Clinical Pharmacology. 2022;88(3):1235–1245.

59. Karlsson I, Boström H. Predicting adverse drug events using heterogeneous event sequences. In: 2016 IEEE International Conference on Healthcare Informatics (ICHI). IEEE; 2016. p. 356–362.

60. Zhao J, Henriksson A, Asker L, Boström H. Predictive modeling of structured electronic health records for adverse drug event detection. BMC medical informatics and decision making. 2015;15:1–15.

61. Mashima Y, Tamura T, Kunikata J, Tada S, Yamada A, Tanigawa M, et al. Using natural language processing techniques to detect adverse events from progress notes due to chemotherapy. Cancer Informatics. 2022;21:11769351221085064.

62. Corny J, Rajkumar A, Martin O, Dode X, Lajonchère JP, Billuart O, et al. A machine learning–based clinical decision support system to identify prescriptions with a high risk of medication error. Journal of the American Medical Informatics Association. 2020;27(11):1688–1694.

63. Udomnuchaisup P, Imsombut A, Suwannahitatorn P, Saethang T. Analysis of the 5Rs in Thailand Medication Error Classification through Natural Language Processing. In: 2023 20th International Joint Conference on Computer Science and Software Engineering (JCSSE). IEEE; 2023. p. 281–284.

64. Hu Q, Tian F, Jin Z, Lin G, Teng F, Xu T. Developing a warning model of potentially inappropriate medications in older Chinese outpatients in tertiary hospitals: a machine-learning study. Journal of Clinical Medicine. 2023;12(7):2619.

65. Yalçin N, Kaşikci M, Çelik HT, Allegaert K, Demirkan K, Yiğit Ş, et al. Development and validation of a machine learning-based detection system to improve precision screening for medication errors in the neonatal intensive care unit. Frontiers in pharmacology. 2023;14:1151560.

66. King CR, Abraham J, Fritz BA, Cui Z, Galanter W, Chen Y, et al. Predicting self-intercepted medication ordering errors using machine learning. PloS one. 2021;16(7):e0254358.

67. Hovor C, O’Donnell LT. Probabilistic risk analysis of medication error. Quality Management in Healthcare. 2007;16(4):349–353.

68. Lo YC, Varghese S, Blackley S, Seger DL, Blumenthal KG, Goss FR, et al. Reconciling allergy information in the electronic health record after a drug challenge using natural language processing. Frontiers in Allergy. 2022;3:904923.

69. Segal G, Segev A, Brom A, Lifshitz Y, Wasserstrum Y, Zimlichman E. Reducing drug prescription errors and adverse drug events by application of a probabilistic, machine-learning based clinical decision support system in an inpatient setting. Journal of the American Medical Informatics Association. 2019;26(12):1560–1565.

70. Schiff GD, Volk LA, Volodarskaya M, Williams DH, Walsh L, Myers SG, et al. Screening for medication errors using an outlier detection system. Journal of the American Medical Informatics Association. 2017;24(2):281–287.

71. Wong ZSY. Statistical classification of drug incidents due to look-alike sound-alike mix-ups. Health informatics journal. 2016;22(2):276–292.

72. Rozenblum R, Rodriguez-Monguio R, Volk LA, Forsythe KJ, Myers S, McGurrin M, et al. Using a machine learning system to identify and prevent medication prescribing errors: a clinical and cost analysis evaluation. The Joint Commission Journal on Quality and Patient Safety. 2020;46(1):3–10.

73. Pais V, Rao S, Muniyal B. Performance Evaluation of Machine Learning Algorithms to Predict the Medication Prescription Errors in Intensive Care Units. In: 2023 International Conference for Advancement in Technology (ICONAT). IEEE; 2023. p. 1–5.

74. Shin SK, Hur H, Cheon EK, Oh OH, Lee JS, Ko WJ, et al. A Personalized and Learning Approach for Identifying Drugs with Adverse Events. Yonsei Medical Journal. 2017;58(6):1229–1236.

75. Dasgupta S, Jayagopal A, Hong ALJ, Mariappan R, Rajan V, et al. Adverse drug event prediction using noisy literature-derived knowledge graphs: algorithm development and validation. JMIR Medical Informatics. 2021;9(10):e32730.

76. Bouzillé G, Morival C, Westerlynck R, Lemordant P, Chazard E, Lecorre P, et al. An automated detection system of drug-drug interactions from electronic patient records using big data analytics. In: MEDINFO 2019: Health and Wellbeing e-Networks for All. IOS Press; 2019. p. 45–49.

77. Rebane J, Karlsson I, Papapetrou P. An investigation of interpretable deep learning for adverse drug event prediction. In: 2019 IEEE 32nd International Symposium on Computer-Based Medical Systems (CBMS). IEEE; 2019. p. 337–342.

78. Rebane J, Samsten I, Pantelidis P, Papapetrou P. Assessing the clinical validity of attention-based and SHAP temporal explanations for adverse drug event predictions. In: 2021 IEEE 34th International Symposium on Computer-Based Medical Systems (CBMS). IEEE; 2021. p. 235–240.

79. Karlsson I, Zhao J. Dimensionality reduction with random indexing: an application on adverse drug event detection using electronic health records. In: 2014 IEEE 27th International Symposium on Computer-Based Medical Systems. IEEE; 2014. p. 304–307.

80. Talukder AK, Selg E, Fernandez R, Raj TD, Waghmare AV, Haas RE. Drugomics: Knowledge Graph & AI to Construct Physicians’ Brain Digital Twin to Prevent Drug Side-Effects and Patient Harm. In: International Conference on Big Data Analytics. Springer; 2022. p. 149–158.

81. Crielaard L, Papapetrou P. Explainable predictions of adverse drug events from electronic health records via oracle coaching. In: 2018 IEEE International Conference on Data Mining Workshops (ICDMW). IEEE; 2018. p. 707–714.

82. Rebane J, Samsten I, Papapetrou P. Exploiting complex medical data with interpretable deep learning for adverse drug event prediction. Artificial Intelligence in Medicine. 2020;109:101942.

83. Karlsson I, Boström H. Handling sparsity with random forests when predicting adverse drug events from electronic health records. In: 2014 ieee international conference on healthcare informatics. IEEE; 2014. p. 17–22.

84. Li J, Ji X, Hua L. Improving the prediction of adverse drug events using feature fusion-based predictive network models. IEEE Access. 2020;8:48812–48821.

85. Langenberger B. Machine learning as a tool to identify inpatients who are not at risk of adverse drug events in a large dataset of a tertiary care hospital in the USA. British Journal of Clinical Pharmacology. 2023;89(12):3523–3538.

86. Zhou F, Uddin S. Mining adverse drug events from patients’ disease histories via a GNN-based subgraph prediction method. In: Proceedings of the 2023 Australasian Computer Science Week; 2023. p. 227–230.

87. Hu Q, Wu B, Wu J, Xu T. Predicting adverse drug events in older inpatients: a machine learning study. International Journal of Clinical Pharmacy. 2022;44(6):1304–1311.

88. Akyon SH, Akyon FC, Yılmaz TE. Artificial intelligence-supported web application design and development for reducing polypharmacy side effects and supporting rational drug use in geriatric patients. Frontiers in Medicine. 2023;10:1029198.

89. Fahmi A, Wong D, Walker L, Buchan I, Pirmohamed M, Sharma A, et al. Combinations of medicines in patients with polypharmacy aged 65–100 in primary care: Large variability in risks of adverse drug related and emergency hospital admissions. Plos one. 2023;18(2):e0281466.

90. Stafford G, Villén N, Roso-Llorach A, Troncoso-Marinõ A, Monteagudo M, Violán C. Combined Multimorbidity and Polypharmacy Patterns in the Elderly: A Cross-Sectional Study in Primary Health Care. International Journal of Environmental Research and Public Health. 2021;18(17):9216. doi:10.3390/ijerph18179216.

91. Lakizadeh A, Babaei M. Detection of polypharmacy side effects by integrating multiple data sources and convolutional neural networks. Molecular Diversity. 2022;26(6):3193–3203.

92. Keine D, Zelek M, Walker JQ, Sabbagh MN. Polypharmacy in an elderly population: enhancing medication management through the use of clinical decision support software platforms. Neurology and therapy. 2019;8:79–94.

93. Seyedtabib M, Kamyari N. Predicting polypharmacy in half a million adults in the Iranian population: comparison of machine learning algorithms. BMC medical informatics and decision making. 2023;23(1):84.

94. Li Q, Spooner SA, Kaiser M, Lingren N, Robbins J, Lingren T, et al. An end-to-end hybrid algorithm for automated medication discrepancy detection. BMC medical informatics and decision making. 2015;15:1–12.

95. Vallamkonda S, Ortega CA, Lo YC, Blackley SV, Wang L, Seger DL, et al. Identifying and reconciling patients’ allergy information within the electronic health record. In: MEDINFO 2021: One World, One Health–Global Partnership for Digital Innovation. IOS Press; 2022. p. 120–124.

96. Cimino JJ, Bright TJ, Li J, et al. Medication reconciliation using natural language processing and controlled terminologies. Studies in health technology and informatics. 2007;129(1):679.

97. Schnipper JL, Nieva HR, Yoon C, Mallouk M, Mixon AS, Rennke S, et al. What works in medication reconciliation: an on-treatment and site analysis of the MARQUIS2 study. BMJ quality & safety. 2023;32(8):457–469.

98. Mouazer A, Leguillon R, Leroy B, Sedki K, Simon C, Falcoff H, et al. ABiMed: towards an innovative clinical decision support system for medication reviews and polypharmacy management. In: Informatics and Technology in Clinical Care and Public Health. IOS Press; 2022. p. 61–64.

99. Millán-Hernández CE, García-Hernández RA, Ledeneva Y. An evolutionary logistic regression method to identify confused drug names. Journal of Intelligent & Fuzzy Systems. 2019;36(5):4609–4619.

100. Levivien C, Cavagna P, Grah A, Buronfosse A, Courseau R, Bézie Y, et al. Assessment of a hybrid decision support system using machine learning with artificial intelligence to safely rule out prescriptions from medication review in daily practice. International Journal of Clinical Pharmacy. 2022;44(2):459–465.

101. Nagata K, Tsuji T, Suetsugu K, Muraoka K, Watanabe H, Kanaya A, et al. Detection of overdose and underdose prescriptions—an unsupervised machine learning approach. PloS one. 2021;16(11):e0260315.

102. Leal Rodríguez C, Haue AD, Mazzoni G, Eriksson R, Hernansanz Biel J, Cantwell L, et al. Drug dosage modifications in 24 million in-patient prescriptions covering eight years: A Danish population-wide study of polypharmacy. PLOS digital health. 2023;2(9):e0000336.

103. Bao Y, Jiang X. An intelligent medicine recommender system framework. In: 2016 IEEE 11Th conference on industrial electronics and applications (ICIEA). IEEE; 2016. p. 1383–1388.

104. Ghasemi SH, Etminani K, Dehghan H, Eslami S, Hasibian MR, Vakili Arki H, et al. Design and evaluation of a smart medication recommendation system for the electronic prescription. In: dHealth 2019–From eHealth to dHealth. IOS Press; 2019. p. 128–135.

105. Hasan S, Duncan GT, Neill DB, Padman R. Towards a collaborative filtering approach to medication reconciliation. In: AMIA Annual Symposium Proceedings. vol. 2008. American Medical Informatics Association; 2008. p. 288.

106. Bergkvist A, Midlöv P, Höglund P, Larsson L, Bondesson Å, Eriksson T. Improved quality in the hospital discharge summary reduces medication errors—LIMM: Landskrona Integrated Medicines Management. European journal of clinical pharmacology. 2009;65:1037–1046.

107. Clyne B, Bradley MC, Smith SM, Hughes CM, Motterlini N, Clear D, et al. Effectiveness of medicines review with web-based pharmaceutical treatment algorithms in reducing potentially inappropriate prescribing in older people in primary care: a cluster randomized trial (OPTI-SCRIPT study protocol). Trials. 2013;14:1–12.

108. Hu G, Yu B, Doctor D. PharmBERT: a Fine-tuned Model for Pharmaceutical Error Prediction. In: 2023 IEEE Conference on Artificial Intelligence (CAI). IEEE; 2023. p. 343–344.

109. Keller MS, Qureshi N, Albertson E, Pevnick J, Brandt N, Bui A, et al. Comparing risk prediction models aimed at predicting hospitalizations for adverse drug events in community dwelling older adults: a protocol paper. Research Square. 2023;.

110. Tursunalieva A, Alexander DL, Dunne R, Li J, Riera L, Zhao Y. Making Sense of Machine Learning: A Review of Interpretation Techniques and Their Applications. Applied Sciences. 2024;14(2):496.

111. Sadeghi Z, Alizadehsani R, Cifci MA, Kausar S, Rehman R, Mahanta P, et al. A review of Explainable Artificial Intelligence in healthcare. Computers and Electrical Engineering. 2024;118:109370.

112. Shahmoradi L, Safdari R, Ahmadi H, Zahmatkeshan M. Clinical decision support systems-based interventions to improve medication outcomes: a systematic literature review on features and effects. Medical Journal of the Islamic Republic of Iran. 2021;35:27.

113. Elbeddini A, Almasalkhi S, Prabaharan T, Tran C, Gazarin M, Elshahawi A. Avoiding a Med-Wreck: a structured medication reconciliation framework and standardized auditing tool utilized to optimize patient safety and reallocate hospital resources. Journal of Pharmaceutical Policy and Practice. 2021;14(1):10.

114. Syrowatka A, Motala A, Lawson E, Shekelle P. Computerized Clinical Decision Support To Prevent Medication Errors and Adverse Drug Events: Rapid Review. Making Healthcare Safer IV: A Continuous Updating of Patient Safety Harms and Practices [Internet]. 2023;.

115. DrFirst. Medication History for Reconciliation: Optimize Workflows with Clinical AI; 2024. Available from: https://drfirst.com/challenges/medication-history-for-reconciliation/.

116. Cureatr. Comprehensive Medication Management Technology; 2024. Available from: https://www.cureatr.com/comprehensive-medication-management-technology.

117. Avicenna Medical Systems. VA Inpatient Medication Reconciliation and Clinical Workflow Software; 2024. Available from: https://www.avicenna-medical.com/va-inpatient-medrec-clinical-workflow-software.

118. HCS, Inc. HCS Medication Reconciliation; 2024. Available from: http://www.hcsinc.net/products/hcs-medication-reconciliation.html.

119. MEDITECH. MEDITECH Expanse Pharmacy; 2024. Available from: https://ehr.meditech.com/ehr-solutions/expanse-pharmacy.

120. WellSky. Ensure patient safety and enhance profitability with WellSky Medication Management; 2024. Available from: https://wellsky.com/medication-management/.

121. Better. Better Meds features at a glance; 2024. Available from: https://www.better.care/better-meds/.

122. (FDB) FD. Medication Reconciliation; 2024. Available from: https://www.fdbhealth.com/applications/medication-reconciliation.

123. RXNT. Electronic Prescribing Software; 2024. Available from: https://www.rxnt.com/software/electronic-prescribing/.

124. Albahri AS, Duhaim AM, Fadhel MA, Alnoor A, Baqer NS, Alzubaidi L, et al. A systematic review of trustworthy and explainable artificial intelligence in healthcare: Assessment of quality, bias risk, and data fusion. Information Fusion. 2023;96:156–191.

125. Kim MK, Rouphael C, McMichael J, Welch N, Dasarathy S. Challenges in and Opportunities for Electronic Health Record-Based Data Analysis and Interpretation. Gut and Liver. 2024;18(2):201.

126. Baardman L, Cristian R, Perakis G, Singhvi D, Skali Lami O, Thayaparan L. The role of optimization in some recent advances in data-driven decision-making. Mathematical Programming. 2023;200(1):1–35.

127. Imrie F, Davis R, van der Schaar M. Multiple stakeholders drive diverse interpretability requirements for machine learning in healthcare. Nature Machine Intelligence. 2023;5(8):824–829.

128. Assiri GA, Shebl NA, Mahmoud MA, Aloudah N, Grant E, Aljadhey H, et al. What is the epidemiology of medication errors, error-related adverse events and risk factors for errors in adults managed in community care contexts? A systematic review of the international literature. BMJ open. 2018;8(5):e019101.

129. Langenberg C, Hingorani AD, Whitty CJ. Biological and functional multimorbidity—from mechanisms to management. Nature Medicine. 2023;29(7):1649–1657.

130. Schmitz K, Lenssen R, Rückbeil M, Berning D, Thomeczek C, Brokmann JC, et al. The WHO High 5s project: medication reconciliation in a German university hospital. A prospective observational cohort study. Zeitschrift für Evidenz, Fortbildung und Qualität im Gesundheitswesen. 2022;168:27–32.

131. Kamekis A, Bertsias A, Moschandreas J, Petelos E, Papadakaki M, Tsiantou V, et al. Patients’ intention to consume prescribed and non-prescribed medicines: A study based on the theory of planned behaviour in selected European countries. Journal of clinical pharmacy and therapeutics. 2018;43(1):26–35.

132. Joseph AL, Borycki EM, Kushniruk AW. Alert fatigue and errors caused by technology: a scoping review and introduction to the flow of cognitive processing model. Knowledge management & e-learning. 2021;13(4):500.

133. Marien S, Krug B, Spinewine A. Electronic tools to support medication reconciliation: a systematic review. Journal of the American Medical Informatics Association. 2017;24(1):227–240.

134. Tamblyn R, Winslade N, Lee TC, Motulsky A, Meguerditchian A, Bustillo M, et al. Improving patient safety and efficiency of medication reconciliation through the development and adoption of a computer-assisted tool with automated electronic integration of population-based community drug data: the RightRx project. Journal of the American Medical Informatics Association. 2018;25(5):482–495.

